# Sex Specific Genomic Insights into Type 1 Diabetes through GWAS and Single Cell Transcriptome Analysis

**DOI:** 10.1101/2025.07.27.25332270

**Authors:** Hui-Qi Qu, Kayleigh Ostberg, Diana J Slater, Fengxiang Wang, James Snyder, Cuiping Hou, John J Connolly, Michael March, Joseph T Glessner, Charlly Kao, Hakon Hakonarson

## Abstract

**Background:** Type 1 diabetes (T1D) exhibits sex differences in genetic risk, yet most genetic studies treat sex as a covariate rather than a potential modifier of risk. We hypothesized that sex-stratified genome-wide association studies (GWAS) would uncover sex specific genetic architecture and improve risk prediction for T1D.

**Methods:** We performed GWAS in 6,599 T1D cases (3,483 males, 3,109 females, 7 undetermined) and 12,350 controls (6,665 males, 5,658 females, 27 undetermined) of European ancestry, testing both additive and additive-by-sex interaction models. We then conducted GWAS separately in males and females. For mechanistic insights into sex-specific effects, we generated single-cell RNA-sequencing (scRNA-seq) profiles of peripheral blood mononuclear cells (PBMCs) from nine matched male-female pediatric pairs of European ancestry. Finally, we tested male-, female-, and standard (all-samples) polygenic risk scores (PRS) in an independent cohort (471 T1D cases, 2,300 controls), and compared their performance by receiver operating characteristic (ROC) analysis.

**Results:** Sex-stratified analyses identified 215 genome wide significant SNPs (P<5×10^-8^) exhibiting significant heterogeneity between sexes: 119 male-specific, 94 female-specific, and two shared SNPs at HLA-B (rs2249932 and rs2249934). Integration of scRNA-seq data pinpointed 41 genes with sex-specific T1D associations that also showed differential expression between males and females in particular cell types. In the independent cohort, sex specific PRS significantly outperformed the combined PRS: in males, AUC=0.668 versus 0.623 (Δ=0.045; DeLong’s p<2.2×10^-16^); in females, AUC=0.719 versus 0.635 (Δ=0.084; DeLong’s p<2.2×10^-16^).

**Conclusions:** Sex-stratified GWAS reveal novel T1D risk loci influenced by sex. Incorporating sex-specific effect sizes into PRS markedly enhances risk discrimination, underscoring the value of sex-aware genetic analyses for precise prediction and intervention in T1D.

## Introduction

Type 1 diabetes (T1D) is a chronic autoimmune disease marked by T cell–mediated destruction of insulin-producing pancreatic β cells^1^. With a peak incidence in childhood and adolescence, T1D affects roughly 15 per 100,000 children worldwide each year, and its prevalence has been rising steadily over the past decades^2^. Although the strongest genetic risk factors lie within the *HLA* region, genome-wide association studies (GWAS) have identified numerous non-HLA loci that modulate susceptibility^3–5^, implicating diverse pathways in immune regulation, β-cell survival, and cytokine signaling, and enable prediction of T1D risk^6^. Epidemiological data suggest sex differences in T1D. In populations with high T1D incidence (predominantly European-origin), males often exhibit a slightly higher incidence in childhood-onset cohorts, whereas females may experience a more aggressive disease course and greater susceptibility to secondary autoimmune complications^7^. Yet traditional GWAS have combined males and females, potentially obscuring sex-specific genetic effects. Sex-stratified and sex-interaction GWAS can uncover loci whose contributions differ between males and females, insights that may refine risk prediction and illuminate distinct etiological pathways.

To understand sex-specific genetic associations with sex-modified immune-cell function, we leveraged single-cell RNA sequencing (scRNA-seq) of peripheral blood mononuclear cells (PBMCs) from matched male–female pediatric pairs. Sex-biased gene expression is a biologically important phenomenon observed across various species, manifesting in differing expression levels of genes between males and females. These differences can vary significantly among tissues and developmental stages^8^. Transcriptomic profiling for genome-wide comparisons of gene expression has revealed many sex-biased genes^8^, a divergence driven by evolutionary forces such as sexual selection and sexual antagonism^9^. The GTEx project has further illuminated this in humans, showing that 37percent of genes exhibit sex-biased expression in at least one tissue, largely in a tissue-specific manner^10^. Differential expression (DE) genes with tissue-specific sex differences are also recognized^11^. The mechanisms behind sex-biased gene expression are multifaceted, involving genetic, hormonal, and epigenetic factors^10^. Understanding these patterns is clinically significant, as sex differences influence disease susceptibility and treatment outcomes^12^. For instance, autoimmune diseases such as systemic lupus erythematosus (SLE) and multiple sclerosis (MS) are more prevalent in women^13^. Moreover, treatment responses and adverse reactions often vary by sex^14^. In PBMCs, sex-specific expression patterns impact immune responses^15^. Understanding these intricate regulatory networks is essential for comprehending sex effects in inflammatory and autoimmune diseases^13,16^. This may inform the development of precision medicine that considers sex-specific differences in gene expression, thereby improving the efficacy of therapeutic interventions for various diseases^17^.

In this study we mapped the sex-specific genetic architecture of T1D through large-scale GWAS. Our scRNA-seq data provide mechanistic understanding of sex-specific genetic associations. By leveraging sex-stratified GWAS, we delineate how sex shapes immunity in T1D, and improve risk prediction using polygenic risk score (PRS).

## Methods

### GWAS

#### Samples and genotyping

Detailed information is available in our previous publication^18^. This study comprised 18,949 individuals of confirmed European ancestry: 6,599 T1D cases (3,483 males, 3,109 females, and 7 of unknown sex) and 12,350 controls (6,665 males, 5,658 females, and 27 of unknown sex). T1D cases were recruited from the Montreal Children’s Hospital, the Children’s Hospital of Philadelphia (CHOP)^19^, the DCCT-EDIC cohort (dbGaP phs000086.v2.p1), and the Type 1 Diabetes Genetics Consortium (T1DGC; dbGaP phs000180.v1.p1). Written informed consent was obtained from all participants. Genotyping was performed using Illumina BeadChips covering at least 550,000 SNPs, and ancestry was confirmed by principal component analysis (PCA) of genomic markers. Genome-wide imputation was conducted via the TOPMed Imputation Server (Version R2, GRCh38).

#### Statistics

We tested 104,689,647 autosomal SNPs with imputation quality R² ≥ 0.3, using both additive (ADD) and additive-by-sex (Add×Sex) models in PLINK v1.9^20^, adjusting for sex and the first ten principal components (PCs) of population structure. We then conducted sex-stratified genome-wide association studies separately in males and females.

### scRNA-seq

#### Samples and sequencing

The PBMC samples and sequencing were carried out as previously described^21^. In detail, we studied PBMCs from 9 matched pairs of de-identified female and male children. The Institutional Review Board at the Children’s Hospital of Philadelphia (CHOP) approved this study. Blood samples from each individual were collected in EDTA-coated tubes. These samples were promptly processed at the Center for Applied Genomics (CAG) at CHOP to isolate PBMCs using Ficoll density gradient centrifugation. scRNA-seq was performed using the 10X Chromium Single Cell Gene Expression assay (10x Genomics, Single Cell 3’ v3)^22^.

Sequencing was done using the Illumina HiSeq2500 SBS v4. The output data from the Chromium single-cell RNA sequencing were analyzed using the Cell Ranger 7.1.0 software suite (10x Genomics), with sequencing reads mapped to the GRCh38 reference genome.

#### Data analysis

Data analysis was conducted using the Seurat R package, employing SCTransform for data normalization and scaling ^23,24^. To improve comparability across samples, the workflow utilized Harmony^25^ to align datasets after principal component analysis (PCA).

This approach facilitated clustering and visualization of cell populations across different samples. Uniform manifold approximation and projection (UMAP)^26^ was employed for grouping cells into clusters. Cell typing was done using singleR and the celldex::DatabaseImmuneCellExpression Data() function^27^ (Fig.2). DE analysis was done in 15 cell types (Fig.2). A gene with DE was defined as FDR<0.05 in at least 2 pairs of samples, and in the same direction within the same cell type. Using the WebGestalt (WEB-based Gene SeT AnaLysis Toolkit) web tool^28^, over-representation analysis (ORA) of the DE genes was done by the Hallmark gene set collection^29^.

**Figure 1.**
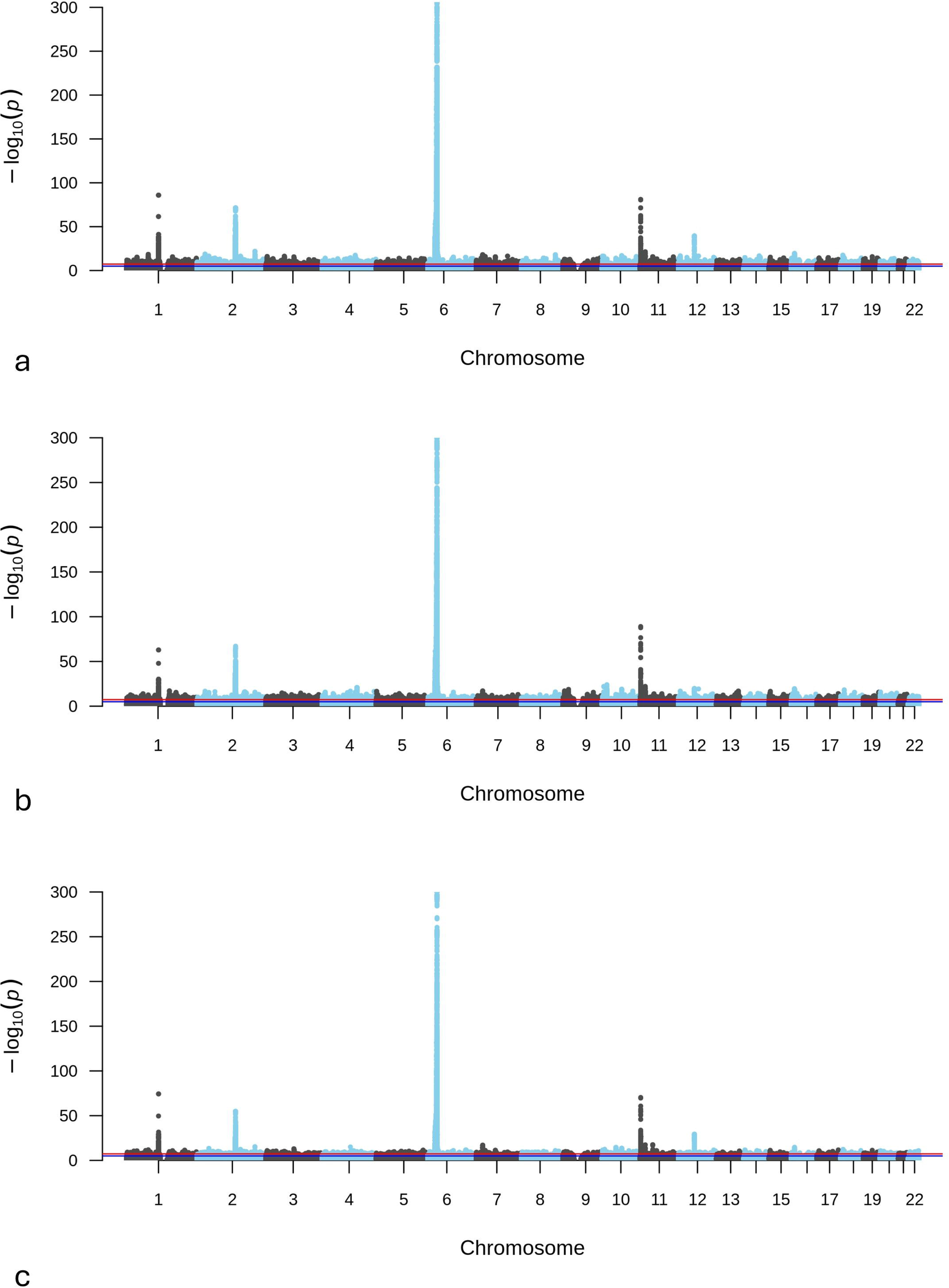
Manhattan plots of genome-wide association results under the additive model for (a) the full cohort, (b) males only, and (c) females only. Each panel displays −log P-values for autosomal SNPs. A red horizontal line denotes the genome-wide significance threshold (P=5×10) and a blue line denotes the suggestive threshold (P=1×10).

**Figure 2.**
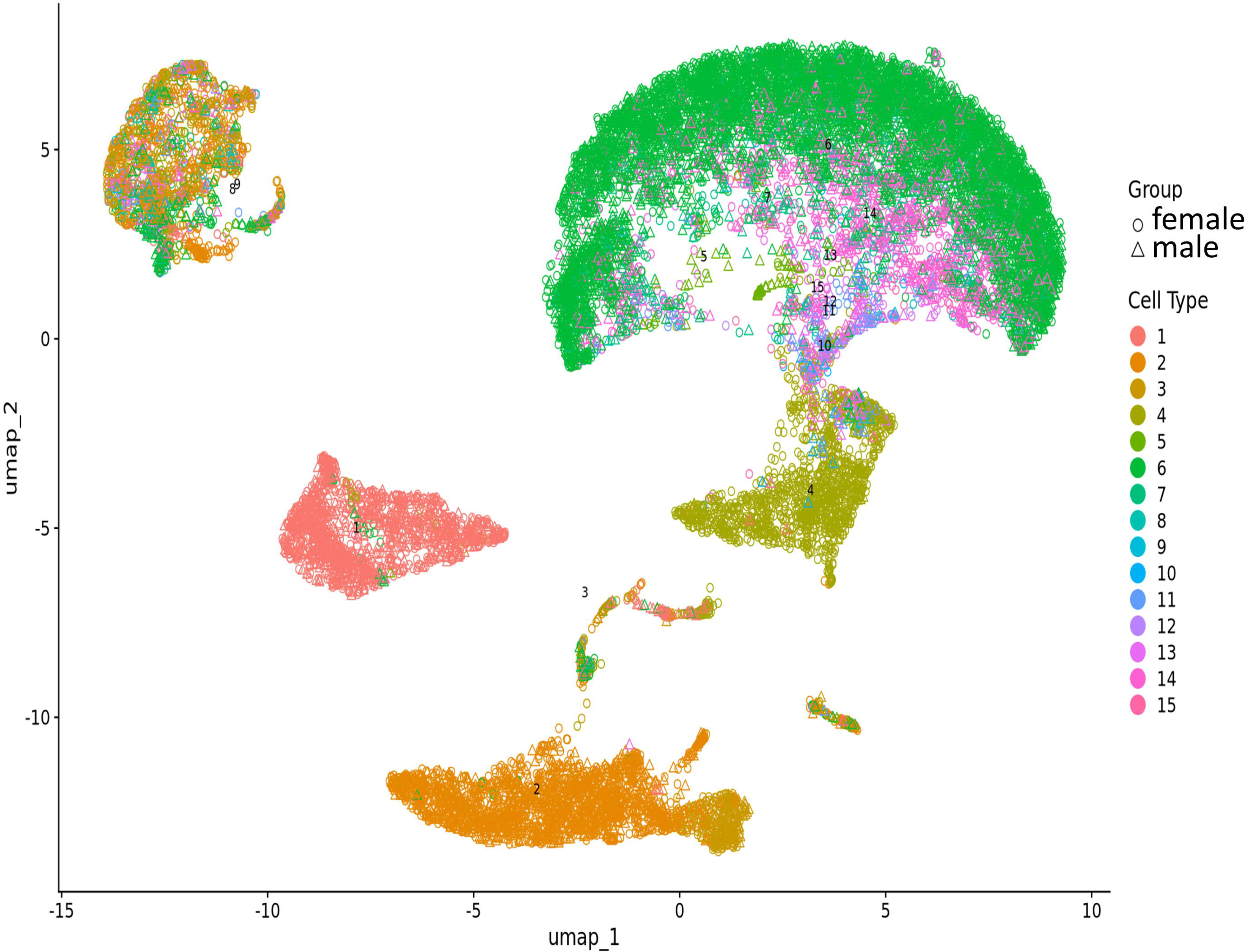
scRNA-seq of PBMCs in female and male patients. Cell types: 1: B cells, naïve; 2: Monocytes, CD14^+^; 3: Monocytes, CD16^+^; 4: NK cells; 5: T cells, CD4^+^, memory TREG; 6: T cells, CD4^+^, naïve; 7: T cells, CD4^+^, naïve TREG; 8: T cells, CD4^+^, naïve, stimulated; 9: T cells, CD4^+^, TFH; 10: T cells, CD4^+^, Th1; 11: T cells, CD4^+^, Th1_17; 12: T cells, CD4^+^, Th17; 13: T cells, CD4^+^, Th2; 14: T cells, CD8^+^, naïve; 15: T cells, CD8^+^, naïve, stimulated.

### Test of sex-specific PRS models in an independent European cohort

#### Samples and genotyping

We evaluated the sex-specific PRS models based on sex-stratified GWAS results in an independent cohort of European ancestry, comprising 471 cases (242 males, 229 females) and 2,300 controls (1,193 males, 1,107 females). All participants were recruited through CHOP, and written informed consent was obtained from all individuals. Genotyping, ancestry confirmation and imputation were performed as described above for the GWAS.

#### PRS construction and evaluation

We LD-clumped SNPs in PLINKv1.9 (1Mb window, 50-SNP step, r²>0.2). Scoring files for the male-specific, female-specific, and combined SNP sets are being deposited in the PGS Catalog. Their assigned identifiers will be provided upon acceptance of the manuscript. We extracted the effect allele and weight for each retained SNP with p-value < 1×10^-4^ from the sex-stratified GWAS results and generated PRS for every individual using PLINK’s scoring function. Separate scores were calculated from the male-specific, female-specific, and all-samples GWAS SNP sets. Discrimination of the standard (all-samples) versus sex-specific PRS was compared within each sex by constructing ROC curves and estimating 95 % confidence intervals via bootstrap resampling. Paired ROC comparisons were conducted using both DeLong’s test and a bootstrap-based test to assess whether the sex-specific PRS provided significantly different discrimination. ROC curves and paired comparisons were generated with the pROC R package^30^.

## Results

### GWAS loci

In the Add×Sex interaction analysis, 2,126 SNPs across 441 gene regions (MAF>0.001) reached P<1×10^-4^ (Supplementary Table1). Of these, 915 SNPs had P > 0.05 under the standard additive model and therefore would not be included in a traditional PRS model. However, all 2,126 SNPs showed nominal significance (P < 0.05) in either the male or female sex-specific association tests. Specifically, 602 SNPs had P < 1×10^-4^ in males, 432 had P < 1×10^-4^ in females, and 12 had P < 1×10^-4^ in both sexes (Fig.1). At the genome-wide significance threshold (P < 5×10^-8^), 215 SNPs were significant in at least one sex:

- Shared Signals (*HLA-B* locus): Two perfectly linked SNPs (chr6:31359384:T>C [rs2249932] and chr6:31359388:T>C [rs2249934], r² = 1) reached genome-wide significance in both sexes. The minor alleles were protective in both males and females, but the effect size was significantly stronger in males (i.e., lower ORs; heterogeneity p = 0.0002).
- Male-Specific Signals: 119 SNPs reached P < 5×10^-8^ in males. (1) Of these, 21 SNPs were also nominally significant in females (P < 0.05). In all 21 SNPs, the minor alleles were protective in both sexes, with significantly stronger effects in males. (2) The remaining 98 SNPs were not significant in females (P > 0.05); among these, 48 SNPs showed opposite effect directions between sexes.
- Female-Specific Signals: 94 SNPs reached P<5×10^-8^ in females. None of these SNPs were significant in males (minimum P = 0.131).

The GWAS summary statistics are available from the NHGRI-EBI GWAS Catalog (GCP001356).

### scRNA-seq results

#### DE genes

A total of 2649 genes in different cell types (Supplementary Table 2) showed significant DE in the same direction in at least two independent pairs of samples (Supplementary Table 3). The cell-specific expression of these genes is depicted in Fig.3. A number of gene sets are significantly overrepresented (Table 1). In addition to DE genes shared by different cell types, a substantial number of DE genes are exclusive to specific cell types (Fig.3D, Supplementary Table 4). Several gene sets are over-represented by these cell type-exclusive genes. In monocytes, genes involved in the interferon (IFN) γ/α responses are exclusively overrepresented. In B cells, genes involved in mTORC1 signaling, unfolded protein response [endoplasmic reticulum (ER) stress], and protein secretion are exclusively overrepresented. In CD8^+^ T cells, genes involved in MYC targets, variant 1, are exclusively overrepresented (Table 1).

**Figure 3.**
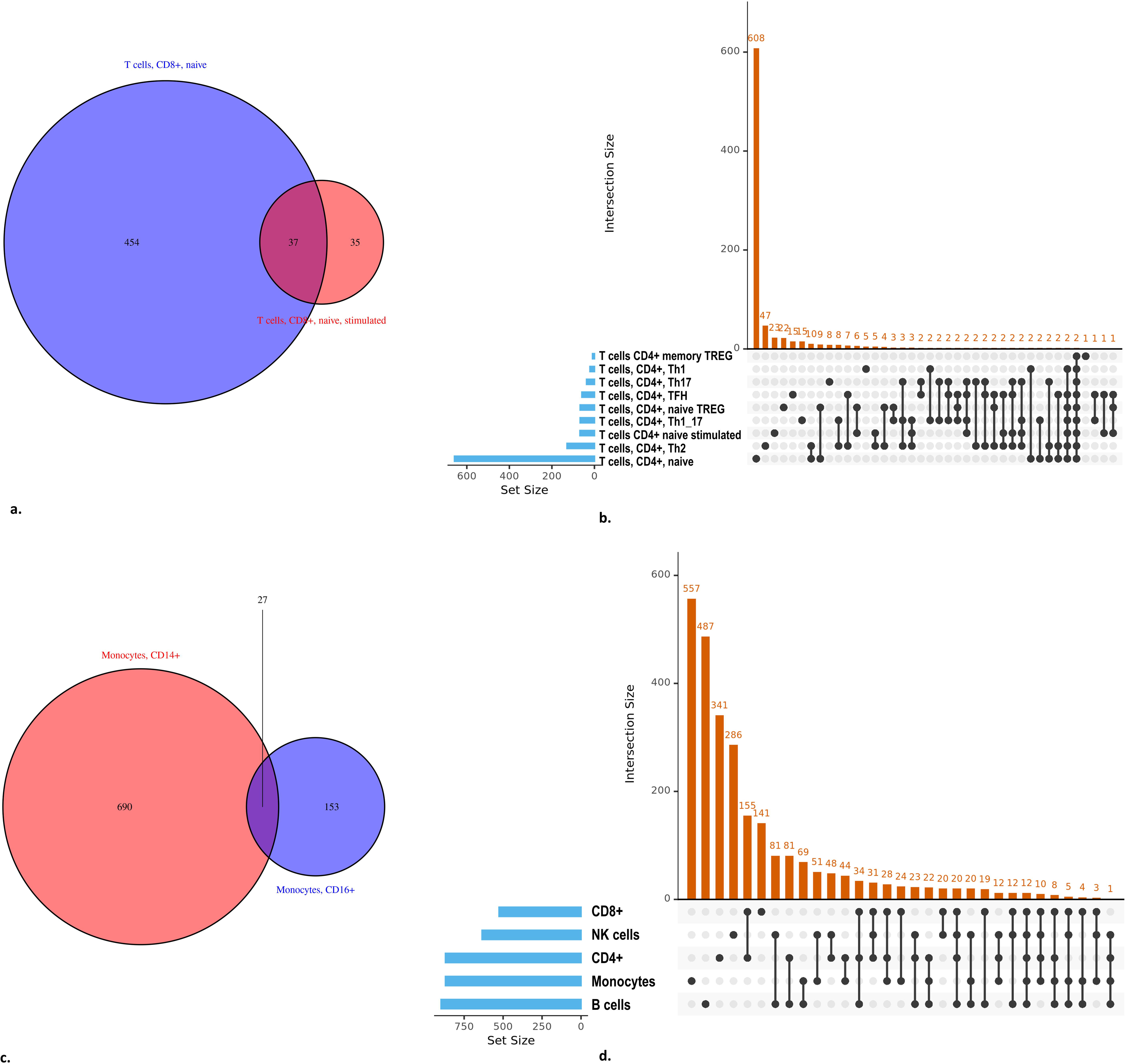
Cell-specific expression of DE genes. a. DE genes in subtypes of CD8^+^ T cells; b. DE genes in subtypes of CD4^+^ T cells; c. DE genes in subtypes of monocytes; d. DE genes in five cell populations.

**Table 1.** Gene sets over-represented by the DE genes.

| Gene Set | Description | Size | Expect | Ratio | P Value | FDR |
| --- | --- | --- | --- | --- | --- | --- |
| **DE genes (n=2649)** |  |  |  |  |  |  |
| HALLMARK_MYC_TARGETS_V1 | MYC targets, variant 1 | 200 | 41.62 | 2.07 | 3.03E-13 | 1.52E-11 |
| HALLMARK_OXIDATIVE_PHOSPHORYLATION | oxidative phosphorylation and citric acid cycle | 200 | 41.62 | 1.80 | 2.08E-08 | 5.20E-07 |
| HALLMARK_INTERFERON_GAMMA_RESPONSE | interferon gamma response | 200 | 41.62 | 1.78 | 5.02E-08 | 8.36E-07 |
| HALLMARK_TNFA_SIGNALING_VIA_NFKB | TNFA signaling via NF-kB | 200 | 41.62 | 1.63 | 6.20E-06 | 7.76E-05 |
| HALLMARK_PROTEIN_SECRETION | protein secretion | 96 | 19.98 | 1.90 | 1.71E-05 | 1.71E-04 |
| HALLMARK_INTERFERON_ALPHA_RESPONSE | interferon alpha response | 97 | 20.18 | 1.83 | 5.73E-05 | 4.77E-04 |
| HALLMARK_PI3K_AKT_MTOR_SIGNALING | PI3K signaling via AKT to mTORC1 | 105 | 21.85 | 1.79 | 7.25E-05 | 5.18E-04 |
| HALLMARK_MTORC1_SIGNALING | mTORC1 signaling | 200 | 41.62 | 1.51 | 1.84E-04 | 1.15E-03 |
| HALLMARK_ANDROGEN_RESPONSE | androgen response | 101 | 21.02 | 1.62 | 1.62E-03 | 9.01E-03 |
| HALLMARK_APOPTOSIS | programmed cell death; caspase pathway | 161 | 33.50 | 1.46 | 2.16E-03 | 1.08E-02 |
| **monocytes (number of exclusive genes 557)** |  |  |  |  |  |  |
| HALLMARK_INTERFERON_GAMMA_RESPONSE | interferon gamma response | 200 | 9.18 | 3.05 | 6.77E-08 | 3.38E-06 |
| HALLMARK_INTERFERON_ALPHA_RESPONSE | interferon alpha response | 97 | 4.45 | 3.59 | 6.31E-06 | 1.58E-04 |
| **B cells naive (number of exclusive genes 487)** |  |  |  |  |  |  |
| HALLMARK_MTORC1_SIGNALING | mTORC1 signaling | 200 | 6.08 | 2.47 | 9.50E-04 | 0.039 |
| HALLMARK_UNFOLDED_PROTEIN_RESPONSE | unfolded protein response; ER stress | 113 | 3.43 | 2.91 | 2.07E-03 | 0.039 |
| HALLMARK_PROTEIN_SECRETION | protein secretion | 96 | 2.92 | 3.09 | 2.32E-03 | 0.039 |
| **CD8 (number of exclusive genes 141)** |  |  |  |  |  |  |
| HALLMARK_MYC_TARGETS_V1 | MYC targets, variant 1 | 200 | 1.83 | 7.11 | 1.04E-08 | 5.19E-07 |

#### Under-expression of chrX genes in females

Among the DE genes, 74 reside on the chrX. Interestingly, chrX genes (*AKAP17A, ALAS2, BEX3, CD99, CSF2RA, DHRSX, IGBP1, IL3RA, MED14, MORF4L2, OTUD5, PDK3, PIM2, PLCXD1, PLP2, RBMX, RLIM, RPGR, SCML1, SH3KBP1, SLC9A7*) show lower expression in females compared to males. Notably, *ALAS2* exhibits the most notable lower expression in CD4^+^ T cells and monocytes in females. *DHRSX* has higher expression in CD14^+^ monocytes but lower expression in NK cells in females.

Conversely, *MED14* shows higher expression in NK cells and lower expression in naive CD8^+^ T cells in females.

#### Autosomal genes with bidirectional DE

In addition to the 2 chrX genes that exhibit bidirectional DE directions in different cell types, 151 autosomal genes and 2 mitochondrial genes (*MT-ND5* and *MT-ND6*) also show bidirectional DE in different cell types. Specifically, genes involved in Epstein-Barr virus (EBV) infection are significantly overrepresented (FDR=0.031), including *CDK6, HLA-F, IFNAR2, NFKB2, PDIA3, PSMC4, TNFAIP3, TRAF3*, and *VIM*. The cell-type specific expression of these genes involved in EBV infection is illustrated in Fig.4.

**Figure 4.**
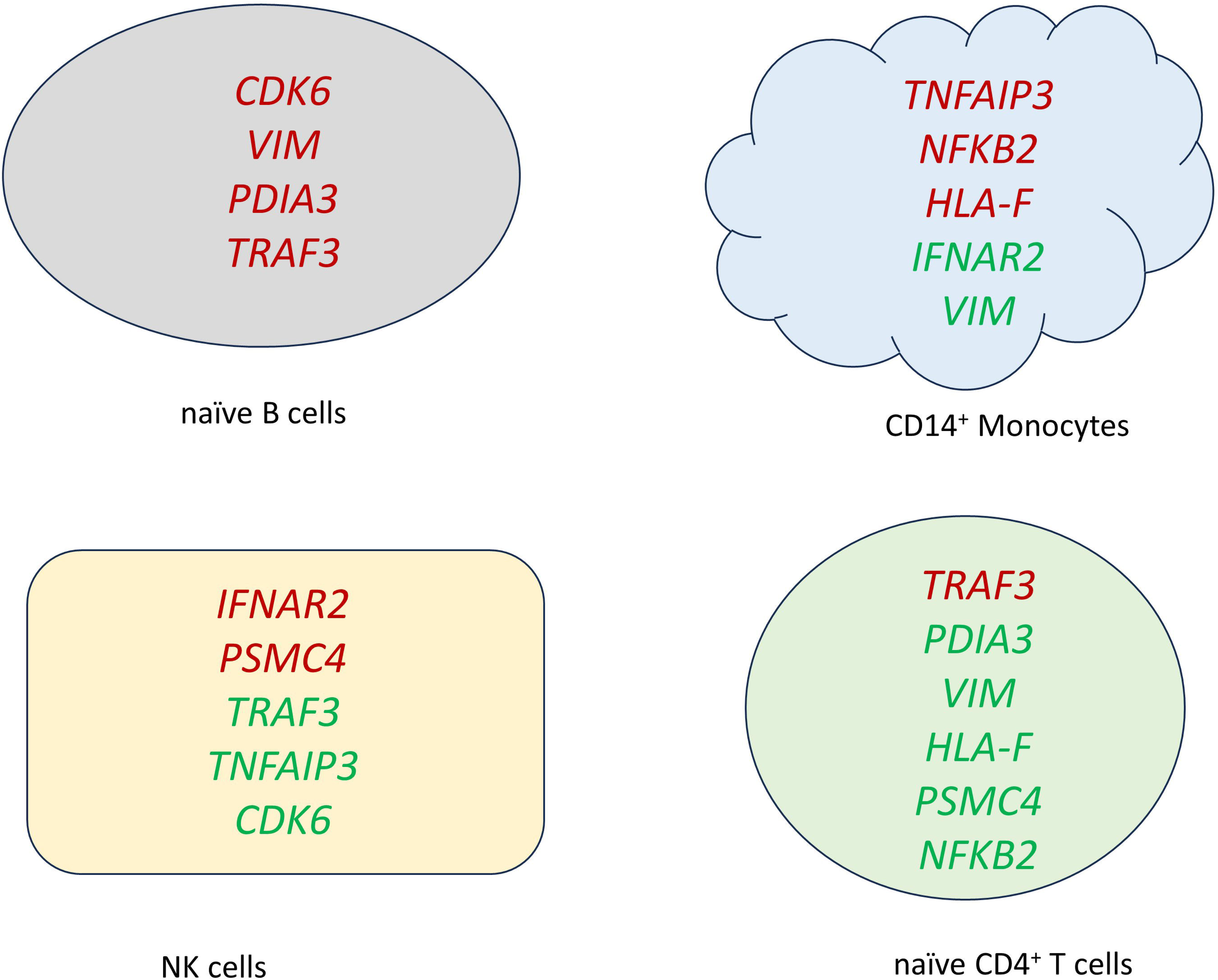
Cell-specific expression of the bidirectional DE genes involved in EBV infection. Genes in red color are down-regulated in females; genes in green color are up-regulated in female**s.**

**Figure 5.**
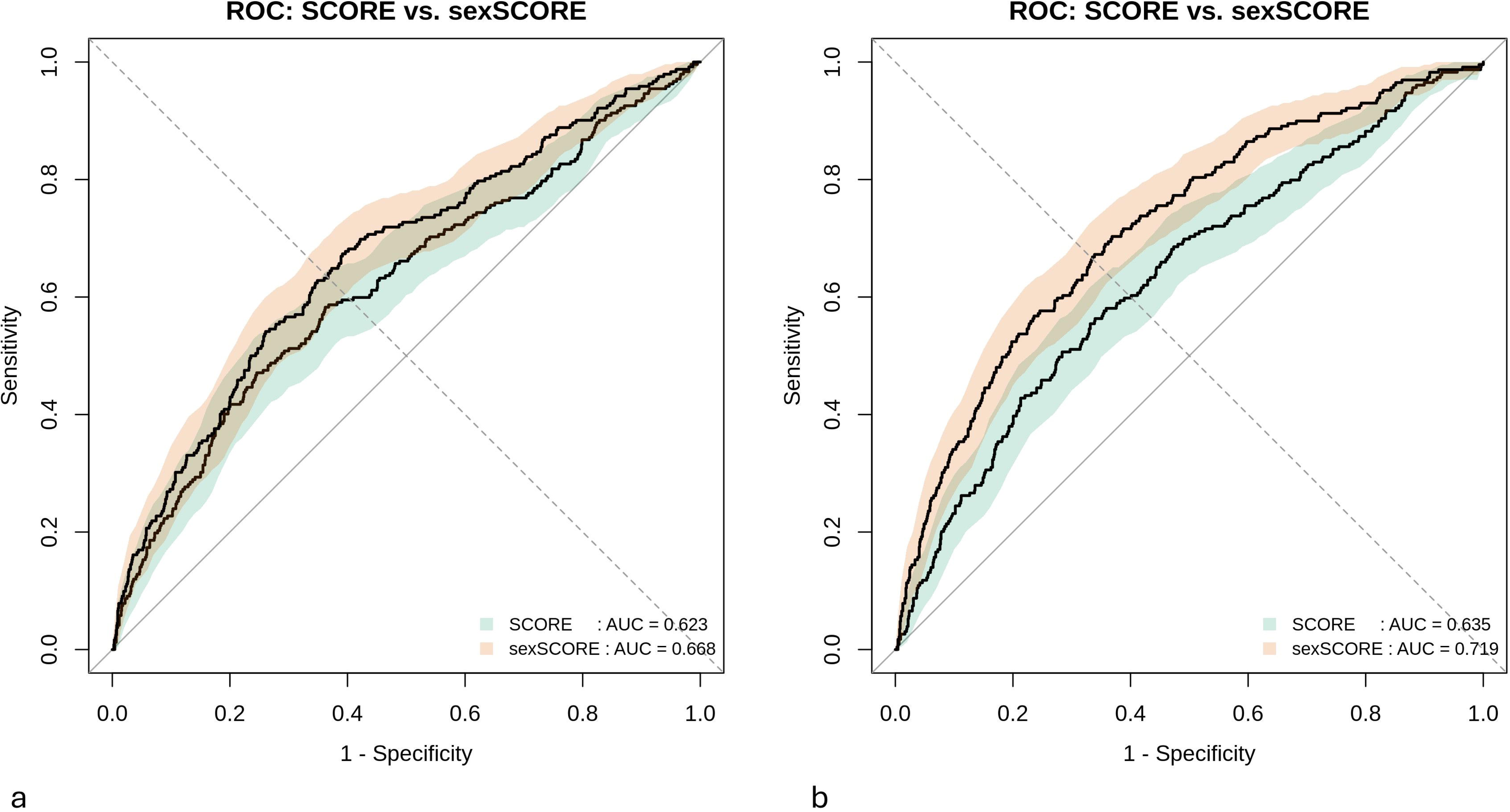
ROC curves comparing the all-sample PRS model (teal) and the sex-specific PRS model (burnt-orange) in (a) male and (b) female participants. Solid curves represent each model’s ROC curve, with semi-transparent shading indicating the 95% confidence interval.

#### Integration of GWAS and scRNA-seq data

Among the loci identified through the Add×Sex interaction analysis, 200 SNPs mapping to 41 genes showed DE in single-cell data (Supplementary Table 5). Of these SNPs,

- 139 SNPs had P < 1×10^-4^ for association with T1D in males
- 17 SNPs had P < 1×10^-4^ for association with T1D in females
- 37 SNPs had P < 0.05 for association with T1D in both sexes. Notably, all exhibited opposite directions of association with T1D between males and females. The genes involved include *ABL2, AGAP1, DOCK5, FLNB, HIP1, MPRIP, NEDD9, PIK3IP1, PISD, PPIP5K2, PXK, RASGRP3, SFI1, SRRM1, SUCLG2, VCP*, and *ZFAND3*.

#### Test of sex-specific PRS in the independent cohort

In males (Fig.5a), the standard (all-samples) PRS achieved an AUC of 0.623 (95% CI, 0.582– 0.664). By contrast, the male-specific PRS yielded a significantly higher AUC of 0.668 (95% CI, 0.629–0.707). A paired comparison of the two correlated ROC curves using DeLong’s test demonstrated a highly significant improvement with the sex-specific score (Z = –8.4547, p < 2.2×10^-16^; ΔAUC = 0.0444, 95% CI [0.0341–0.0546]). A bootstrap-based test (2,000 stratified replicates) confirmed these findings (D = –8.3838, p < 2.2×10^-16^).

In females (Fig.5b), the all-samples PRS produced an AUC of 0.635 (95% CI, 0.594–0.675), whereas the female-specific PRS attained an AUC of 0.719 (95% CI, 0.682–0.756). DeLong’s test indicated a highly significant increase in discrimination for the sex-specific model (Z = – 8.4667, p < 2.2×10^-16^; ΔAUC = 0.0840, 95% CI [0.0646–0.1035]). Bootstrap validation (2,000 stratified replicates) yielded consistent results (D = –8.3857, p < 2.2×10^-16^).

## Discussion

### Insights by sex-specific GWAS

By explicitly modeling the additive-by-sex interaction, we uncovered a substantially expanded set of loci that would have been partially or entirely missed under a standard additive framework. Notably, 915 of these SNPs showed no evidence of association in the pooled (sex-agnostic) analysis (P>0.05), highlighting novel, sex-modulated signals^31,32^.

Among the loci with genome-wide significance, two perfectly linked SNPs in the *HLA-B* locus (rs2249932/rs2249934) achieved genome-wide significance (P<5×10^-8^) in both sexes, yet their protective effect was significantly stronger in males. This finding aligns with the pivotal role of *HLA-B* in antigen presentation and its tight linkage to T1D susceptibility^6^. In particular, *HLA-B*39:06* is known to increase T1D risk by directing CD8^+^ T-cell peptide recognition and promoting autoreactivity^33,34^. Our lead SNPs rs2249932/rs2249934 are in modest LD (r² = 0.301) with the established *HLA-B*39:06* tag SNP rs2844603, suggesting they may partly capture the influence of this high-risk allele. Supporting a sex-dependent context, transcriptomic and proteomic profiling of human islets has revealed that female donors display higher basal expression of *HLA-B* and greater resilience to endoplasmic reticulum stress. Under experimental challenge, using thapsigargin, female islets maintain glucose-stimulated insulin secretion, whereas male islets exhibit pronounced secretory failure^35,36^. Our study shows the minor allele is protective. According to the GTEx (https://www.gtexportal.org/home/snp/rs2249932), the minor allele is correlated with higher *HLA-B* expression in whole blood. In males, who have lower baseline *HLA-B* levels and reduced endoplasmic reticulum stress resilience, this upregulation may bolster antigen presentation dynamics or alter the peptide repertoire in a way that enhances islet survival under stress. In females, who already display higher *HLA-B* expression and greater stress resilience, the same allele would yield a smaller incremental benefit.

The 119 male-specific genome-wide significant SNPs are from 27 gene regions (*ADGRL3, AMPH, ANO1, C6orf15, CTNND1, CUBN, ERG, HCG27, LINC01579, LINC01749, LINC02994, LNC-LBCS, MAP3K8, MED13L, MRPS6, KCNE2, MTCO1P31, MYO16, PCDH1, RAP1GDS1, RETNLB, RNA5SP279, RNU6-1133P, RPS24P7, SNX19, STMN3, TJP1, UBBP5*).

Several converging mechanisms are highlighted:

#### Innate immune and cytokine-driven inflammation

Risk variants in *MAP3K8* (*TPL2*) and *RAP1GDS1* amplify pro-inflammatory signaling only in the context of a male hormone milieu. Testosterone has been shown to transcriptionally regulate *TPL2*, higher androgen levels boost *MAP3K8* expression and downstream ERK activation, whereas estrogens in females tend to dampen TLR/MAPK signaling and promote anti-inflammatory IL-10 production^15,37^. Moreover, male innate immune cells exhibit a more pronounced pro-inflammatory cytokine profile via TLR pathways, so variants that augment *MAP3K8* or *RAP1GDS1* signaling may disproportionately drive β-cell–targeted inflammation in men^38^.

#### Cell–cell adhesion and tissue-barrier integrity

Variants in *CTNND1* (p120-catenin) and *TJP1* (ZO-1) impair junctional stability more severely in males because estrogens in females actively reinforce epithelial and endothelial barriers. *In vitro* and *in vivo* studies demonstrate that estrogen (via ERβ) upregulates ZO-1, alleviates ER stress, and suppresses cytokine-driven permeability, buffering the impact of risk alleles in women^39^. In contrast, the absence of this hormonal backup in men leaves adherens and tight junctions vulnerable, unmasking the deleterious effects of CTNND1/TJP1 SNPs on gut and islet-vascular integrity.

#### Long noncoding RNAs and epigenetic regulation

Male-specific lncRNA loci (e.g. *LINC01579, LINC01749, LINC02994, LNC-LBCS*) harbor androgen-responsive elements and are preferentially expressed in male tissues. Sex steroid–driven chromatin profiling shows that many lncRNAs are directly upregulated by androgens and downregulated by estrogens, leading to stronger cis-and trans-regulatory effects in men^40,41^. In women, estrogen-mediated repression of these lncRNAs buffers genotype-dependent dysregulation of downstream immune or β-cell genes, preventing genome-wide significance in female-only analyses.

#### Mitochondrial quality control and β-cell stress responses

Male β-cells rely on a higher baseline expression of mitochondrial proteins but possess weaker antioxidant defenses. Estrogen in females activates the ERα axis of the mitochondrial UPR (UPR^mt^) and enhances ROS clearance, safeguarding mitochondrial proteostasis under stress^42,43^. Testosterone, by contrast, augments mitochondrial biogenesis without the same UPR^mt^ boost^44^, so *MRPS6* variants that impair the mitochondrial ribosome and UPR^mt^ may manifest only in men as increased β-cell apoptosis and impaired insulin secretion

In females, 94 SNPs passed the genome-wide threshold are from 10 gene regions (*FGF12, LINC01122, LRRC30, PLXNC1, RBFOX2, RNU6-75P, SPTBN1, THAP3P1, ZNF503-AS1, ZNF837*), none of which showed nominal effects in males. This clear demarcation highlights female-exclusive genetic mechanisms:

#### Estrogen-enhanced receptor and growth-factor signaling

*FGF12* belongs to the intracellular FGF family, converging on classical FGF/FGFR pathways that are potently upregulated by estrogen in multiple tissues. ERα activation amplifies FGF-mediated ERK/MAPK signaling to promote cell survival and proliferation; by analogy, female-specific *FGF12* variants likely boost pro-survival signals in β-cells under inflammatory stress^45,46^. *PLXNC1*, the plexin-C1 semaphorin receptor, is a direct transcriptional target of ERβ. Estrogen–ERβ complexes induce PLXNC1 expression to shape semaphorin-mediated guidance cues, tuning dendritic and T-cell trafficking and dampening excessive inflammation in female islet microenvironments^47,48^.

#### Alternative-splicing and small-nuclear RNA networks

*RBFOX2* is a master regulator of alternative splicing for genes driving insulin-granule docking (e.g., *STXBP1*, *SNAP25*)^49^. *RBFOX2* physically interacts with ERα and its expression is stabilized by estrogen^50^, suggesting that female β-cells leverage enhanced splicing fidelity under cytokine challenge. *RNU6-75P* represents a U6 snRNA pseudogene; U6 snRNAs are central to the spliceosomal catalytic core, and snRNA abundance and modification are dynamically regulated by hormonal milieu. Female-biased estrogen signaling alters snRNA transcription in immune cells, implying that *RNU6-75P* variants fine-tune spliceosome composition in women with T1D^51^.

#### Female-specific long noncoding and zinc-finger regulatory axes

*LINC01122*, *ZNF503-AS1*, and *THAP3P1* are lncRNAs transcribed from female-specific T1D loci. They harbor estrogen-response elements and scaffold chromatin modifiers to orchestrate transcriptional programs in female β-cells and T-lymphocytes^52^. *ZNF837*, a KRAB-zinc-finger protein, co-occupies many ERα binding sites. Female-specific *ZNF837* variants likely modulate ERα chromatin recruitment at autoimmune loci, reshaping transcriptional networks that protect against β-cell autoimmunity^53^.

#### Cytoskeletal and cell–cell interaction remodeling

SPTBN1 (βII-spectrin) scaffolds the plasma membrane to the actin cytoskeleton, maintaining epithelial and endothelial barrier integrity.

Estrogen upregulates spectrin family members in vascular and reproductive epithelia, buffering the impact of deleterious SPTBN1 alleles in women^54^. LRRC30, a leucine-rich repeat (LRR) protein, is predicted to participate in innate immune receptor complexes. LRR domains mediate pathogen sensing via TLRs, and ERs directly regulate TLR expression and signalling; female-specific LRRC30 variants may fine-tune innate immune activation in T1D^55,56^.

Crucially, 915 SNPs had P > 0.05 under the standard additive (ADD) model and therefore would not be included in a traditional PRS model. Incorporating these sex-modulated SNPs into sex-specific PRSs holds promise to boost predictive accuracy and facilitate personalized risk stratification.

### Insights into sex-specific gene expression by cell types

#### Differential expression genes affected by sex

We identified 2649 genes with DE affected by sex. A number of gene sets are significantly overrepresented by the DE genes (Table 1): (1) *MYC* targets, variant 1: The *MYC* gene is a transcription factor that regulates the expression of numerous genes involved in cell proliferation, growth, metabolism, and apoptosis in a wide variety of cell types including PBMCs^57,58^, e.g., metabolic reprogramming for immune cell activation and function^59^. *MYC* is essential for pancreatic β-cell proliferation and function, with dysregulated *MYC* levels contributing to β-cell apoptosis and diabetes onset^60^. BCG vaccination in T1D patients upregulates *MYC* and its target genes in monocytes and CD4^+^ T cells, enhancing metabolic pathways that may restore immune tolerance and improve glycemic control^61^. (2) Oxidative phosphorylation and citric acid cycle: oxidative phosphorylation and the citric acid cycle ensures that immune cells have sufficient energy^62^. Oxidative stress and mitochondrial dysfunction are key features of T1D, with genes in OXPHOS pathways serving as markers for hyperglycemia-induced oxidative damage in PBMCs and correlating with disease progression^63^. (3) IFN-γ response: These genes related to IFN-γ response play critical roles in the function of antigen-presenting cells^64^, Th1 cell differentiation^65^, cytotoxic T lymphocytes (CTLs)^66^, and cytotoxic activity of NK cells^67^. IFN-γ produced during insulitis synergizes with IL-1β and TNF-α to activate β-cell apoptotic programs, and genetic models show that IFN-γ is essential for β-cell destruction and insulitis in T1D^68^. (4) TNFA signaling via NF-κB: These genes are involved in the activation of various immune cells, promoting inflammation and the immune response to infections^69^. TNF-α together with IL-1β and IFN-γ induces β-cell apoptosis through NF-κB– dependent gene networks, driving the loss of insulin-producing cells in T1D^70^. (5) Protein secretion: Effective protein secretion in PBMCs is essential for immune function, e.g., secreting cytokines and antibodies^71^. Enhanced protein secretion pathways in B cells underpin the production of islet autoantibodies (e.g., against insulin and GAD65), which are predictive markers and mediators of T1D autoimmunity^72^. (6) IFN-α response: These genes play a crucial role by inducing the expression of IFN-stimulated genes (ISGs)^73^ in the activation and function of immune cells such as NK cells and CTLs. Type I interferons, particularly IFN-α, are expressed in islets of at-risk individuals and augment *MHC Class I* expression, promoting autoantigen presentation to autoreactive CD8^+^ T cells in early T1D^74^. (7) PI3K signaling via AKT to mTORC1: It is vital for the activation and differentiation of T cells and B cells^75^.

Modulation of the PI3K/Akt/mTOR pathway with rapamycin and IL-2 in NOD mice limits autoreactive T cell expansion and preserves β-cell mass, highlighting this axis as a therapeutic target in T1D^76^. (8) mTORC1 signaling: It plays a role in regulating the balance between effector T cells and regulatory T cells, impacting immune homeostasis and tolerance^77^. Inhibition of mTORC1 with rapamycin, especially in combination with GABA, ameliorates autoimmune diabetes in NOD mice, demonstrating the critical role of mTORC1 in T1D pathogenesis^78^. (9) Androgen response: In PBMCs, androgens influence the differentiation and activity of various immune cells, including T cells, B cells, and macrophages^79^. More genes involved in androgen response are upregulated in females, whereas several genes have downregulated expression in females in B cells (*ACSL3, CDK6, HMGCS1, RRP12, SEC24D, SLC38A2, ZBTB10*) and monocytes (*ACTN1, B4GALT1, NCOA4, UBE2J1, ZBTB10, ANKH, RRP12, SGK1*).

Endogenous androgens confer protection against T1D in male NOD mice through microbiome-mediated and direct immunomodulatory effects; castration reverses this protection, while 5α-dihydrotestosterone (DHT) supplementation restores it^80,81^. (10) Programmed cell death; caspase pathway: In PBMCs, apoptosis is crucial for immune regulation, ensuring the removal of activated immune cells after an immune response and preventing autoimmunity. Caspase-mediated apoptosis is the main form of β-cell death in T1D, with activated caspase-3 and intrinsic pathway components detected in human islets and contributing to disease progression^82^.

Among these genes, a number of gene sets are over-represented in specific cell-type exclusively. The overrepresentation of genes involved in the IFN-γ and IFN-α responses in monocytes plays important roles in orchestrating the innate immune response against pathogens^83^. IFNs are key cytokines in the activation of macrophages and dendritic cells^83,84^, and monocytes are the circulating precursors of macrophages and dendritic cells^85^. The exclusive overrepresentation of genes associated with mTORC1 signaling, unfolded protein response (ER stress), and protein secretion in B cells play important roles in antibody production and immune regulation. The mTORC1 pathway is essential for B cell development, differentiation, and antibody production^86^. Unfolded protein response reflects the high demand for protein synthesis and secretion during B cell activation and antibody production^87^. The protein secretion pathway is critical for exporting antibodies and cytokines^88^. *MYC* is a transcription factor that regulates various cellular processes, including cell growth, metabolism, and proliferation^89^. The overrepresentation of *MYC* target genes in CD8^+^ T cells involves controlling the metabolic and proliferative activities of T effector functions^90^.

#### Immune implications of underexpressed chrX genes in females

The observed downregulation of chrX genes in females compared to males can be related to X-inactivation^91^ and epigenetic regulation^92^. In particular, the X-chromosome dampening (XCD) mechanism may be involved^93^. For instance, *ALAS2* (5’-aminolevulinate synthase 2) is an enzyme involved in heme biosynthesis^94^. X-inactivation ensures that females do not overexpress *ALAS2*^95^. Heme biosynthesis occurs in most cells of the body, although erythroid cells synthesize most of the total heme of the body^96^. The expression of *ALAS2* in monocytes and CD4^+^ T cells may involve the anti-inflammatory and immunomodulatory functions of heme^97^. Downregulated gene expression in females compared to males affects various blood cell types, leading to significant physiological impacts. In naïve B cells, downregulation of genes like *CD99*, *IGBP1*, and *PIM2* results in impaired cell adhesion^98^, reduced proliferation, and diminished survival^99^. In CD14^+^ monocytes, *OTUD5* affects protein stability, reducing monocyte differentiation and innate immune functionality^100^. CD16^+^ monocytes may have reduced cell signaling and migration due to downregulated genes like *BEX3* and *CD99*. NK cells may have weakened activation and energy supply with downregulation of *PDK3*^101^.

*DHRSX* (dehydrogenase/reductase X-linked) encodes an enzyme belonging to the short-chain dehydrogenase/reductase (SDR) family, involved in oxidation-reduction processes^102^. *MED14* (mediator complex subunit 14) is a critical component of the mediator complex, a transcriptional coactivator^103^. These differential expressions may be attributed to sex-specific regulatory mechanisms, including estrogen influences^104^ with its cell type-specificity^105^.

#### Autosomal genes with cell type-specific bidirectional DE

Sex-specific differences in gene expression within PBMCs are driven by various mechanisms, including hormonal regulation and epigenetic modifications^92^. These include the epigenetic regulation of autosomal gene expression by sex chromosomes^106^. Hormonal influences, particularly from sex hormones like estrogen and testosterone, play a significant role. For example, we observed that the nuclear factor-kappa-B (NF-κB) subunit 2 gene (*NFKB2*) exhibited lower expression in CD14^+^ monocytes but higher expression in naïve CD8^+^ T cells in females. NF-κB signaling regulates the development and survival of immune cells^107,108^. In CD14^+^ monocytes, estrogens may reduce NF-κB pathway activation and suppress *NFKB2* expression^109^. On the other hand, female CD8^+^ T cells have an increased response to IL-12, related to estrogen response or effects of X-linked genes^110^, and IL-12 induces the activation of the NF-κB pathway^111^.

EBV primarily infects B cells. In immunocompromised individuals, T cells and natural killer (NK) cells might also be infected^112^. It can serve as a model for understanding various viral infections. While the basic mechanism of EBV infection is similar in males and females, there are significant differences in immune response, severity of symptoms, and risks of associated diseases between the genders^113^. Among cell type-specific bidirectional DE genes involved in EBV infection, EBV leverages these genes to manipulate host cellular mechanisms, facilitating its replication and enabling it to evade immune detection. By influencing *CDK6*, EBV synchronizes the cell cycle of infected cells to favor viral replication^114^. EBV exploits cellular structures such as vimentin (*VIM*) for the intracellular transport of viral components^115^. EBV manipulates the NF-κB pathway via *NFKB2*, *TNFAIP3*, and *TRAF3* to enhance cell survival and modulate inflammation^116^. *PDIA3* is involved in the proper folding of MHC class I molecules, essential for the presentation of viral antigens on the cell surface^117^. We observed downregulation of CDK6, VIM, PDIA3, and TRAF3 in female naïve B cells, which may contribute to less severe symptoms in some cases but also predisposes women to a higher risk of autoimmune disorders linked to EBV, such as systemic lupus erythematosus (SLE) and multiple sclerosis (MS)^118^.

The scRNA-seq study uncovers a diverse landscape of sex-influenced gene expression programs across PBMC subsets. These insights into cell-type-specific, sex biased transcriptional networks deepen our understanding of immune regulation and T1D pathogenesis, and may inform development of targeted, sex informed therapies for autoimmune and infectious diseases.

### Functional Insights into T1D GWAS Loci from scRNA-seq Data

By Integration of sex-stratified GWAS interaction signals with single-cell transcriptomes of PBMCs, insights can be gained from the 37 SNPs whose minor alleles have opposite effects in males versus females, and whose expression is highly cell-type–specific. This striking reversal of effect may reflect three interlocking mechanisms: (1) hormone-dependent regulation of allele-specific enhancers, whereby estrogen and androgen receptors recruit coactivators or corepressors in a sex-biased manner; (2) sex-specific chromatin landscapes, as male and female immune cells show differential DNA methylation and histone modification profiles that alter transcription factor access; and (3) immune-cell composition differences, since men and women harbor varying proportions of monocyte, T-cell, B-cell and NK-cell subsets, each with its own regulatory milieu^15,119^. At the transcriptional level, large-scale analyses (e.g., GTEx v8) have demonstrated that while sex-biased gene expression is widespread, affecting over 13,000 genes across tissues, these effects are generally modest and highly tissue-and cell type–specific^10^.

Single-cell transcriptomic studies are useful: a recent atlas revealed sex-specific expression programs in multiple immune subsets, including naive CD4^+^ T cells, monocytes, B cells, and NK cells^120^.

In our dataset, monocyte-biased genes such as *PISD* (log_2_FC≈1.53 in monocytes) and *DOCK5* (log_2_FC≈1.53 in monocytes; 2.66 in NK cells) are upregulated in females, implicating enhanced mitochondrial and cytoskeletal remodeling pathways that support robust innate responses^121^.

Monocytes are professional antigen-presenting cells and major cytokine producers in early T1D lesions^122^, and NK cells are critical innate sentinels that shape adaptive autoimmunity^123^. PISD encodes mitochondrial phosphatidylserine decarboxylase, catalyzing the conversion of phosphatidylserine to phosphatidylethanolamine and thereby maintaining inner mitochondrial membrane integrity and bioenergetic capacity^124^. DOCK5, a member of the dedicator of cytokinesis family, functions as a guanine nucleotide exchange factor for Rho GTPases, orchestrating actin-and microtubule-remodeling to enhance cell spreading, migration, and degranulation in innate immune cells^125^. Together, these female-biased transcriptional enrichments point to augmented mitochondrial function and cytoskeletal dynamics that likely underpin the heightened phagocytic and migratory capabilities of female monocytes and NK cells.

Conversely, male-biased upregulation of genes like *FLNB* in B cells (log_2_FC ≈ −1.93) and *NEDD9* in NK cells (log_2_FC ≈ −1.16) may reflect attenuated cytoskeletal dynamics and signaling in male lymphocytes. B cells are the source of pathogenic autoantibodies and antigen presentation to T cells^72^. *FLNB* encodes filamin B, a high-molecular-weight actin-crosslinking protein that organizes cortical actin filaments and scaffolds signaling modules (e.g., Rac1–JNK) to control cell shape, migration, and mechanotransduction in lymphocytes^126,127^. *NEDD9* is a non-catalytic scaffolding protein that localizes to focal adhesions in immune cells, coordinating integrin-and Src-family-kinase signaling, actin remodeling, and immunological synapse assembly critical for NK cell motility and cytotoxic function^128^. These male-biased expression shifts in key cytoskeletal regulators suggest that male lymphocytes may have reduced capacity for dynamic actin remodeling and signal propagation, which could contribute to sex differences in immune cell trafficking and effector activity.

### Validation of Sex-Specific PRSs

The aforementioned sex GWAS and sex single-cell RNA-seq results justified testing sex-specific PRSs. To assess the translational utility of our sex-aware modeling framework, we evaluated sex-specific PRSs in an independent cohort. In males, the sex-specific score achieved an AUC of 0.668 versus 0.623 for the standard PRS (ΔAUC = 0.0444; DeLong’s Z = –8.4547, p < 2.2×10^-16^), with bootstrap validation confirming these gains. In females, the female-specific PRS reached an AUC of 0.719 compared to 0.635 for the all-samples model (ΔAUC = 0.0840; Z = – 8.4667, p < 2.2×10^-16^). The larger ΔAUC in females likely reflects both stronger effect sizes at sex-interaction GWAS loci. By capturing this sex-dependent genetic architecture, our PRSs yield more accurate risk stratification, paving the way for sex-informed screening and prevention strategies in T1D. By incorporating the GRS2 algorithm and markers, this is a significant improvement^129^. Nevertheless, our cohort is predominantly of European ancestry, and performance may vary in other populations; future work should extend these analyses to diverse ancestries, integrate additional molecular annotations (e.g., chromatin accessibility), and evaluate prospective clinical utility.

In conclusion, by integrating additive-by-sex GWAS interactions, we have uncovered sex-modulated genetic loci. Cell-type–specific expression programs, ranging from hormone-dependent enhancers and chromatin differences to immune-composition biases, provides reasonable explanation of the sex effects. These efforts enabled the construction of sex-specific PRSs that significantly outperform standard models in independent cohorts, illustrating the power of single-cell resolution and sex-aware design for precision risk stratification and paving the way toward tailored, sex-informed strategies for T1D prediction and intervention.

## Data Availability

The GWAS summary statistics are available from the NHGRI-EBI GWAS Catalog (GCP001356). Scoring files for the male-specific, female-specific, and combined SNP sets will be deposited in the PGS Catalog upon assignment of a DOI for this submission. Their PGS Catalog identifiers will be provided in an update to the medRxiv preprint. Additional information is available from the corresponding author upon request.

## Acknowledgements

We thank all patients and their families who have participated in our research for the past two decades.

## Ethics approval and consent to participate

All experimental protocols were approved by the Institutional Review Board (IRB) of the Children’s Hospital of Philadelphia (CHOP) with the IRB number: IRB 16-013278. Informed consent was obtained from all subjects. If subjects are under 18, consent was also obtained from a parent and/or legal guardian with assent from the child if 7 years or older.

## Consent for publication

Not applicable.

## Supplementary Tables

**Supplementary Table 1** GWAS-Identified Loci (including all samples, interaction test, and sex-stratified analyses)

**Supplementary Table 2** Cell counts of each cell type in each sample

**Supplementary Table 3** All genes with differential expression in at least two independent sample pairs (Log2FC values). Positive log2FC indicates higher expression in females.

**Supplementary Table 4** Genes with cell-type-specific DE

**Supplementary Table 5** 200 SNPs from 41 genes with additive-by-sex interactions show differential expression in single-cell data.

